# Effect of nasal carriage of *Bacillus* species on COVID-19 severity: A cross-sectional study

**DOI:** 10.1101/2023.04.15.23288553

**Authors:** Muinah A. Fowora, Adenike Aiyedogbon, Ibilola Omolopo, Ahmed O. Tajudeen, Abdul-Lateef Olanlege, Adefunke Abioye, Grace B. Akintunde, Morenike O. Folayan, Babatunde L. Salako

## Abstract

Intranasal sprays containing *Bacillus* species are being researched for treating viral respiratory tract infections. The aim of this study was to assess the relationship between the nasal carriage of *Bacillus* and COVID-19 severity. This was a cross-sectional study that collected nasopharyngeal samples from adults 18 years and above visiting two COVID-19 testing centers in Lagos, Nigeria between September 2020 and September 2021. *Bacillus* species were cultured from the respiratory samples and confirmed using molecular methods. The dependent variable was COVID-19 status classified as negative, asymptomatic, mild, or severe. The independent variable was the nasal carriage of *Bacillus* species. Multinomial regression analysis was done to determine the association between nasal carriage of *Bacillus* and COVID-19 severity after adjusting for age, sex, and co-morbidity status. About 388 participants were included in the study with a mean (standard deviation) age of 40.05 (13.563) years. The majority (61.1%) of the participants were male, 100 (25.8%) had severe COVID-19, 130 (33.5%) had pre-existing comorbidity, and 76 (19.6%) had *Bacillus* cultured from their nasopharyngeal specimen. Bacillus species presence was significantly associated with higher odds of severe COVID-19 compared to having a negative COVID-19 status. However, the presence of *Bacillus* species was significantly associated with lower odds of severe COVID-19 compared to having a mild COVID-19 status. The study suggests that nasal carriage of *Bacillus* species may substantially impact the clinical course of COVID-19. This study supports the exploration of *Bacillus* species in the prevention and management of viral respiratory tract infections.

**IMPORTANCE:** With the introduction of intranasal spray containing *Bacillus* species for the treatment of viral respiratory tract infections, such as COVID-19 and respiratory syncytial virus, identifying the association between the nasal carriage of *Bacillus* species and COVID-19 susceptibility and severity will help further substantiate the investigation of these bacteria for COVID-19 prevention and treatment. This study evaluated the association between the carriage of *Bacillus* species in the nasopharyngeal tract and COVID-19 severity and found that the presence of *Bacillus* species in the nasopharynx may significantly impact the clinical course of COVID-19.

## BACKGROUND

Corona Virus Disease 2019 (COVID-19), an acute and highly infectious respiratory disease caused by the severe acute respiratory syndrome coronavirus 2 (SARS-CoV-2) has led to the death of more than 6.42 million people globally since its outbreak in Wuhan, China in 2019 [1]. Several factors such as the nasal microbiota determine the susceptibility and severity of the disease [2-7]. Microbiota can confer some level of protection on the host against some diseases by creating a unique microbial ecosystem that enhances resistance against the manifestation of respiratory tract infection caused by bacterial, fungal, and/or viral pathogens [8, 9]. It also serves as markers of disease [10] and the regulation of local and systemic immunity in the nasopharyngeal tract thereby influencing COVID-19 susceptibility and clinical outcome [7]. In addition, viral infection modifies the host’s nasal microbiota, decreases the nasal and gut microbiota diversity, and increases the disease infectivity and severity [11, 12].

A significant increase in bacteria belonging to the phyla Bacteroides, Proteobacteria, Actinobacteria, Firmicutes, and Fusobacteria have been reported among COVID-19 patients [13] though some other study did not find a significant difference in the nasal bacterial composition and diversity in patients with COVID-19 when compared with patients negative for COVID-19 [14]. The members of bacteria belonging to the phylum Firmicutes, especially of the class Bacilli, have been associated with respiratory immunity and strengthening the host defense against respiratory viral infections [11]. *Bacillus* has been explored as intranasal sprays for the treatment of viral infection [15, 16] as it eases the symptoms of acute respiratory tract infection caused by respiratory syncytial virus by reducing the viral load and inflammation associated with the disease [16]. *Bacillus subtilis* possesses the probiotic potential to increase mucosal and tonsillar immunity against respiratory diseases through the formation of immune cells in the nasal cavity and tonsil [15]. Like bacilli intranasal sprays, lactobacilli-containing throat sprays have also been demonstrated to considerably lessen the acute symptoms of COVID-19 as well as the load of the virus in the nasopharynx [17]. However, there are currently no studies showing the effect of nasal carriage of *Bacillus* species on the susceptibility to SARS-CoV-2 infection and severity.

The aim of this study was to determine the association between nasal carriage of *Bacillus species* and COVID-19 susceptibility and severity. We hypothesize that there will be a negative association between nasal carriage of *Bacillus* and COVID-19 susceptibility and severity.

## METHODS

### Study design and population

This study was a cross-sectional study that compared the prevalence of nasal carriage of *Bacillus* species among adult who were tested positive or negative for COVID-19 in Lagos State, Nigeria. The study population included individuals ≥ 18 years, who present for SARS-CoV-2 testing at the COVID-19 modified drive-through centre of the Nigerian Institute of Medical Research and the laboratory of the Infectious Disease Hospital, Lagos. The participants were recruited between September 2020 to September 2021. Adults who had travelled out of the country in the last one month of the survey were excluded from the study. Informed consents were appropriately obtained from all eligible participants.

### Sample size and sampling technique

The sample size for this study was determined using the formula for sample size calculations for prevalence studies [18]. The standard normal variate was set at 5% type I error; the precision was set at 0.05; and the expected proportion of adults with nasal *Bacillus* was set at 50% in the absence of any available data. The minimum sample size was 385. The sample size was increased to 450 participants to account for missing data or invalid COVID-19 results. A convenient sample of participants willing to participate in the study was recruited.

### Data collection

Information on the age (age as at last birthday) and sex (sex at birth) of participants were collected. Other information collected were the symptoms and severity of the patient and if the participant had any underlying conditions such as diabetes, asthma, hypertension, heart, lung, and kidney disease. These variables had been identified factors that influence COVID-19 susceptibility and severity [19, 20]. No identifying information was collected from the participants.

Participants were classified as positive for COVID-19 when the results on the real-time reverse transcription–polymerase-chain-reaction (RT-PCR) assay of their nasopharyngeal swab specimens was positive. Participants with positive test results were categorized as asymptomatic, mild or severe cases using the guidelines of the Nigeria Centre for Disease Control and the National Institute of Health [21, 22]. Asymptomatic cases were defined as patients that had no symptoms specific to COVID-19 but tested positive SARS-CoV-2. Mild cases were patients who had symptoms corresponding to COVID-19 such as fever (<38^0^C), cough, sore throat, malaise, headache, muscle pain, loss of taste and smell, nausea, vomiting, diarrhea, abdominal pain, but do not have difficulty breathing, dyspnea and any underlying conditions. Severe cases were patients with clinical manifestations of COVID-19 in conjunction with difficulty in breathing, reduced/decreased breath sounds, dullness in percussion, increased or decreased vocal resonance, and presence of an underlying comorbid condition such as diabetes, asthma, hypertension, heart, lung, asthma and kidney disease.

### Sample collection and analysis

Nasopharyngeal swab samples were collected from the participants into two different transport media. One containing a viral transport medium (VTM) and the other a bacterial transport medium containing Tryptone soy broth buffered with 2.5% saline and 10% glycerol. All samples were analyzed within one to two hours of specimen collection.

#### RNA extraction and qRT-PCR

Viral Nucleic acid Extraction was carried out on the nasopharyngeal sample in VTM using the Qiagen Viral Extraction kit according to manufacturer’s instructions, and SARS-CoV-2 was detected by quantitative reverse transcription polymerase chain reaction (qRT-PCR) using a commercial Nucleic Acid Diagnostic Kit for COVID-19, Sansure Novel Coronavirus (2019-nCoV), according to manufacturer’s protocol (Sansure Biotech Inc). The Sansure kit has high sensitivity and is endorsed for the SARS-CoV-2 testing [23].

#### Bacteria isolation

The nasopharyngeal samples in the bacteria transport medium were plated on Mueller Hinton Agar and de Man-Rogosa Sharpe (MRS) agar (Oxoid, USA) using a disposable loop. The nutrient agar was incubated aerobically at 37^0^C for 24 hours while the MRS agar was incubated at 37^0^C under anaerobic conditions using a anaerobic jar containing an anaerobic gas generating pack (Thermo Scientific™ Oxoid™ AnaeroGen™) for 48 to 72 hours.

#### Bacterial identification

Isolates were identified based on the morphological characteristics of the colony and using both phenotypic and molecular methods. The phenotypic based methods included Gram staining, nitrate reduction, gelatin hydrolysis, and citrate utilization as previously described [24, 25].

Deoxyribonucleic acid (DNA) was extracted from all the presumptively identified bacteria using the NIMR Biotech DNA extraction kit. To confirm the identification of the *Bacillus* species, polymerase chain reaction amplification of the 16s rRNA gene specific for *Bacillus* was carried out. The reaction was done using the primer set 16S-HV (Forward primer -5’-TGTAAAACGACGGCCAGTGCCTAATACATGCAAGTCGAGCG-3’ and Reverse primer-5’-CAGGAAACAGCTATGACCACTGCTGCCTCCCGTAGGAGT-3’) as previously described [26].

Polymerase chain reaction was done using the Solis Biodyne 5X HOT FIREPol Blend Master mix in a 20 μl reaction mixture, containing 1X Blend Master mix buffer (Solis Biodyne), 1.5 mM MgCl2, 200μM of each deoxynucleoside triphosphates (dNTP)(Solis Biodyne), 25pMol of each primer (BIOMERS, Germany), 2 unit of Hot FIREPol DNA polymerase (Solis Biodyne), Proofreading Enzyme, 5μl of the extracted DNA, and sterile nuclease-free water was used to make up the reaction mixture.

Thermal cycling was conducted in a PTC 200 gradient thermal Cycler Eppendorf for an initial denaturation of 95°C for 15 minutes followed by 35 amplification cycles of 30 seconds at 95°C, one minute at 58°C and one minute 30 Seconds at 72°C. This was followed by a final extension step of 10 minutes at 72°C. The amplification product was separated on a 1.5% agarose gel and electrophoresis was carried out at 80V for 1 hour 30 minutes. After electrophoresis, DNA bands were visualized by ethidium bromide staining. 100bp DNA ladder was used as DNA molecular weight standard.

## Data Availability

All data produced in the present study are available upon reasonable request to the authors.

## STATISTICAL ANALYSIS

The study variables were presented as means (standard deviations) or frequencies and percentages. A multivariate logistic regression model was developed to determine the association between COVID-19 status (negative, asymptomatic, mild and severe) and *Bacillus* species isolation status dichotomized into positive and negative. The model was adjusted for age, sex, and presence of comorbidity. Statistical analysis was done using SPSS version 26.0 and statistical significance was set at a p value < 0.05.

## ETHICAL CONSIDERATIONS

Ethical approval was obtained from the Institutional Review Board of the Nigerian Institute of Medical Research (IRB/20/043). A written informed consent was obtained from all the participants following the national ethics guidelines [27].

## RESULTS

There were 450 study participants recruited into this study. Forty-three (9.6%) had invalid COVID-19 qRT-PCR and the information on the age and/or sex was missing for 19 (4.2%) participants. The complete data for 388 participants is presented in Table 1.

**Table 1.** Age, sex, COVID-19 severity, pre-existing conditions, and *Bacillus* isolation from the nasopharyngeal samples of adult Nigerians with COVID-19 (N = 388)

| Factor | Variable | Statistic |
| --- | --- | --- |
| Sex | Female: n (%) | 151 (38.9) |
|  | Male: n (%) | 237 (61.1) |
| Age | Mean (SD) | 40.05 (13.563) |
| COVID-19 | Negative n (%) | 228 (58.8) |
|  | Asymptomatic n (%) | 28 (7.2) |
|  | Mild n (%) | 32 (8.2) |
|  | Severe n (%) | 100 (25.8) |
| Pre-existing Condition | Yes n (%) | 130 (33.5) |
|  | No n (%) | 258 (66.5) |
| Type of pre-existing condition | None n (%) | 258 (66.5) |
|  | Asthma n (%) | 15 (3.9) |
|  | Diabetes n (%) | 14 (3.6) |
|  | Hepatitis B n (%) | 3 (0.8) |
|  | High blood pressure/Hypertension n (%) | 71 (18.3) |
|  | Rhinosinusitis n (%) | 1 (0.3) |
|  | Diabetes and High Blood Pressure n (%) | 22 (5.7) |
|  | Ulcer n (%) | 4 (1.0) |
| *Bacillus* isolated | No n (%) | 312 (80.4%) |
|  | Yes n (%) | 76 (19.6) |

The age of participants ranged from 18 to 75 years with the mean age (standard deviation) age of 40.05 (13.563) years. Also, 237 (61.1%) participants were male, 228 (58.8%) had a negative COVID-19 test result and 258 (66.5%) had no pre-existing medical condition. Among participants that reported having a pre-existing condition, 71 (18.3%) had high blood pressure and 22 (5.7%) had both type II diabetes mellitus and high blood pressure. There were *Bacillus* species isolated from 76 (19.6%) of the nasopharyngeal samples collected.

Table 2 shows that the presence of *Bacillus* species in the nasopharyngeal samples was also associated with COVID-19 severity: patients with *Bacillus* species in the nasopharyngeal samples had significantly higher odds for severe COVID-19 than a negative COVID-19 result (AOR = 3.347, 95% CI: 1.359, 8.243). Also, patients with *Bacillus* species in the nasopharyngeal samples had significantly lower odds for severe COVID-19 than a mild COVID-19 result (AOR = 0.158, 95% CI: 0.055, 0.455).

**Table 2.**
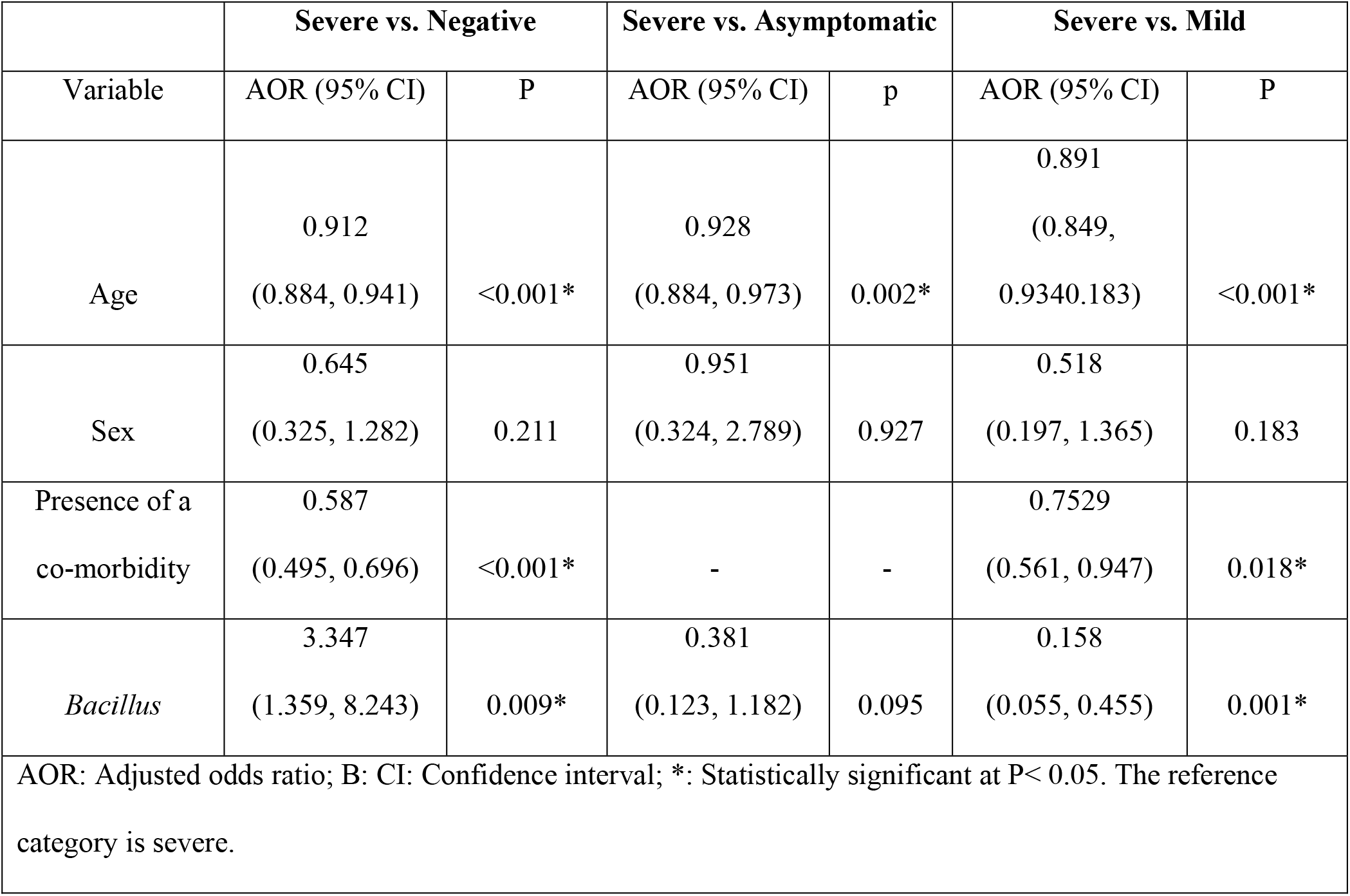
Associations between Age, comorbidity, and the presence of nasal carriage of *Bacillus* species and COVID-19 Severity (N = 388).

## Discussion

The objective of this study was to determine if there was an association between the nasal carriage of *Bacillus* species and COVID-19 severity. The study findings showed a strong positive association between nasal carriage of *Bacillus* species and COVID-19 severity while participants who carried *Bacillus* species in their upper respiratory tract had a lower likelihood of developing severe COVID-19 when compared with mild cases. The study findings did not support the study hypothesis of a negative relationship between nasal carriage of *Bacillus* species and COVID-19 severity.

This is the first study showing a relationship between bacteria co-infection involving nasal carriage of *Bacillus* species and COVID-19. The *Bacillus* species isolated in this study can be classified as a co-infection as they were recovered from the participants at the point of SARS-CoV-2 infection diagnosis [37]. Our study results indicated that attention needs to be given to *Bacillus* species as a secondary bacterial co-infection in COVID-19 and a possible risk factor for severe COVID 19. Prior studies that evaluated respiratory bacteria co-infection in patients with COVID-19 identified *Acinetobacter baumannii, Staphylococcus aureus, Klebsiella pneumoniae* and *Staphylococcus aureus* as the most isolated bacterial infection from respiratory tract cultures [38-40]. We had also reported an association between the nasal carriage of *Moraxella catarrhalis* and *Chlamydophila pneumoniae* and COVID-19 [41].

The observed association between *Bacillus species*, a respiratory opportunistic pathogen, and the severity of COVID-19 may be connected to its association with the high risk for co-morbidities in patients with severe COVID-19. *Bacillus* counts are high in patients who are immunocompromised and patients with diabetes [28, 30-32]. These co-morbidities increase the risk for severe COVID-19. The study finding may influence the development of nasal sprays for the treatment of respiratory viral infections [15, 16]. There is still a need to further identify (up to strain level) the *Bacillus* species isolated and characterize the isolates by antibiotic susceptibility and virulence to better understand the properties of the isolates that may influence the severity of COVID-19.

We also found that individuals with *Bacillus* species in their upper respiratory tract may have a lower likelihood of developing severe COVID-19 when compared with mild cases. This finding is similar to the outcome of a prior study that demonstrated that treatment with lactobacilli had the potential to lower the viral load of SARs-COV-2 in the upper respiratory tract without necessarily resolving the symptoms of COVID-19 [17]. The lower likelihood of severe COVID-19 compared to mild COVID-19 when *Bacillus* is present may also be related to the reduction or improvement of the symptoms of SARs-COV-2 infection by the *Bacillus* species. Metabolites, such as surfactins, iturin A, and fengycins produced by some *Bacillus* species have antibacterial and antifungal activities [33-36]. These metabolites may be responsible in easing the symptoms of COVID-19 and preventing the progression of the severity of the disease in mild cases of COVID-19. There is a need to further substantiate this result in a prospective study.

This study has its limitations. The cross-sectional study design used limits the generalizability of the results and can only suggest an association and not a cause-effect relationship. The use of nasopharyngeal respiratory tract cultures also limits the interpretation of this findings as upper respiratory tract samples are more subject to contamination and commensal pathogens, and thus bacterial isolation from such specimen are often higher than those from blood cultures [38]. We could not collect specimen from the lower respiratory tract of the participants in this study as most of the participants did not have a productive cough. However, there is a need for further study to determine the pathogenicity of the *Bacillus* species and the role this plays in COVID-19 severity. The use of lower respiratory tract specimen and/or blood culture should also be explored to further validate the association between *Bacillus* species co-infection and COVID-19 severity.

## CONCLUSION

This study suggests that the presence of *Bacillus* species in the nasopharyngeal tract could significantly influence the clinical progression or severity of COVID-19. This study supports the exploration of *Bacillus* species in the prevention and management of COVID-19.

## ACKNOWLEDGMENTS

This study was funded with funds from the Nigeria Institute of Medical Research (NMG-CIF-18-0036) and the early career research grant of the Royal Society of Tropical Medicine and Hygiene.

We also thank Rukayat Raji and Khadijah Sanusi for their laboratory support, and the Lagos State Ministry of Health.

